# Quantifying the current and future risk of West Nile virus and Usutu virus in humans and horses across England and Wales using an R_0_-based suitability metric

**DOI:** 10.64898/2026.10.04.26364722

**Authors:** Joanna N. de Klerk, Jack Pilgrim, Matthew Baylis, Marcus Blagrove, Jolyon M. Medlock, Alexander G.C. Vaux, Robert C. Bruce, Arran J. Folly, Luigi Sedda

## Abstract

In England and Wales, Usutu virus (USUV) was detected in 2020 and is now established. West Nile virus (WNV) has so far only been detected in a single location, in two *Aedes vexans* mosquito pools sampled during 2023 surveillance. Given the growing threat to the UK from *Culex*-borne viruses there is a need to undertake risk assessments to contribute to preparedness and surveillance strategy for both viruses across human and animal health sectors.

Here, we present the first application of a mechanistic R₀-based model, incorporating temperature dependence and combining data on vector species distribution with projected future climate scenarios, to assess transmission suitability for WNV and USUV across England and Wales.

Under current climate conditions, the two viruses showed comparable spatial patterns, with risk concentrated most heavily in the southeast along the Thames estuary and extending into Kent, Essex and Surrey, as well as around Cambridgeshire and adjacent counties, and across Yorkshire, Nottinghamshire and Lincolnshire. This pattern broadly corresponded with the cumulative distribution of *Culex* mosquito abundance established in our earlier work (*Culex pipiens pipiens, Culex pipiens molestus* and *Culex torrentium*). Both viruses were projected to show increasing transmission suitability risk over time, with substantially steeper increases under Representative Common Pathway (RCP) 8.5 then RCP 4.5. Equine WNV cases were predicted to be most likely in southeast England, in particular around Newmarket, driven by the region’s dense horse population. Among the human population, London was identified as the highest-risk area, with Liverpool and Manchester also showing elevated risk levels, driven by a combination of high human and mosquito distributions.

When human population distributions were age-adjusted to account for the proportion of over 60 years-olds, there was elevated risk in coastal regions, particularly in the east (Norfolk and Suffolk) and southeast (Kent) of England.

The geographic distribution of positive mosquito and bird USUV cases recorded in 2023 and 2024 broadly overlapped with areas predicted to have higher USUV suitability, providing validation for the model’s spatial outputs.

By characterising relative spatial transmission suitability risk, this work adds to a growing understanding of where arbovirus emergence and spread are most likely across England and Wales, and can support decisions around resource allocation, targeted surveillance efforts, and public health readiness. The results also underscore the importance of incorporating climate change and vector ecology into future modelling work, given their likely influence on how arbovirus risk evolves over time.

## Introduction

West Nile virus (WNV) and Usutu virus (USUV) are biologically similar mosquito-borne orthoflaviviruses which are zoonotic emerging threats, with WNV being more significant for public health. WNV transmission to both animals and humans has been recorded in Europe, the Middle East, Africa, India, Australia, North America, Central America, and the Caribbean [1], whereas USUV remains limited to Africa, Europe [2] and the Middle East [3].

Originating in Africa, both viruses were most likely introduced into Europe via migratory birds. They persist in sylvatic transmission cycles between birds and mosquitoes, with occasional spillover into dead-end hosts such as humans and horses, usually causing asymptomatic or mild conditions, although WNV can cause potentially fatal neurological diseases in around 1-2% of the infected people [1]. There is no licenced vaccine available for humans, but there are several for horses. WNV is a notifiable disease in horses, with around 20% developing clinical disease and up to a 30% case fatality rate in those exhibiting symptoms [4]. Nevertheless, the likelihood of clinically severe manifestations is significantly lower in USUV infections than in those caused by WNV [5].

WNV has been found in over 65 mosquito species globally [6], and there is considerable overlap between competent vector species of WNV and USUV. In Europe, *Culex pipiens* sensu lato (*Cx. pipiens* s.l.) Linnaeus, 1758, is the principal vector of both viruses [7,8], although the two *Culex pipiens* s.l. biotypes, *Cx. pipiens pipiens* and *Cx. pipiens molestus,* differ in competence, behaviour, habitat and feeding preferences [9,10]. Over 326 avian species have been implicated in the maintenance of WNV and/or USUV [11], and susceptibility to infection varies among bird species between the two viruses. For USUV, Eurasian Blackbird (*Turdus merula*) populations have undergone large die-offs, and Great grey owls (*Strix nebulosa*) have exhibited high mortality in zoos [12,13], and this has also been the case in the UK. In the case of WNV, members of the crow family (*Corvidae*) have suffered substantial declines in North America; however, in Europe, Africa, and Asia, avian populations have not displayed significant mortality [14].

Within Europe, *Passeriformes* are the amplifying bird hosts of both WNV and USUV [15], and WNV outbreaks appear to be correlated with the flyway of passerines, as well as areas of high bird densities [16]. Birds travelling along the East Atlantic Flyway migrate from northern or western Africa, through the Iberian Peninsula, across France and onwards to the United Kingdom, Ireland or further north [17]. An entry risk assessment by Bessell *et al.* (2016) [17] predicted the probability that WNV would reach the UK during a single whole migratory season was 0.881, if a bird became infected in the Camargue region of France along its route. However, the limiting factor to establishment in the UK was the short remaining infectious period (2.17 days) and declining viraemia, and therefore the probability of onward transmission remained very low. Furthermore, onward transmission also depends on infected birds arriving at a time that coincides with sufficient mosquito abundance and activity to enable local amplification [18]. In the past decade, WNV has expanded northward in Europe, reaching Germany in 2018 [19] and the Netherlands in 2020 [20]. This expansion has created a larger endemic area closer to the UK, driven by changes in temperature, precipitation, and intensified agricultural activity [16,21]. It has raised concerns that migratory birds infected in this region could arrive in the UK during peak viraemia and remain infectious longer, thereby increasing the likelihood of onward transmission.

The emergence of USUV into Austria, Germany, Switzerland and the Netherlands preceded the emergence of WNV by one to seven years [14], and given the similarity of host ranges, competent vector species, and transmission cycles, as well as their frequent co-circulation [22], the presence of USUV may be considered indicative of ecological conditions suitable for WNV circulation, although it does not necessarily predict imminent emergence. In 2020, USUV was first detected in the UK and has subsequently been associated with a 50% decline in Blackbirds in the Greater London area [23,24]. USUV is now regarded as endemic in southern England, having successfully overwintered and been detected annually to date [24,25,26]. In comparison, West Nile virus has not yet become established in the UK, having been detected only once in two pools of *Aedes vexans* mosquitoes during 2023 surveillance in Nottinghamshire, with no subsequent detections in mosquitoes nor birds in the years following [27].

Human USUV infection is most commonly asymptomatic, but neurological cases have been documented in Europe [28]. As of September 2025, no symptomatic or asymptomatic, autochthonous, human cases of USUV have been detected in the UK [29]. Comparatively, there is greater concern over the potential impact WNV could have if it were to establish, as WNV is more likely to cause clinical disease in humans. The UK is an ageing population, with 19.3% over the age of 65; higher than the global mean [30]. Around 2% of WNV infected people over the age of 65 develop neuroinvasive disease that is potentially fatal, which is much higher than the reported 0.1% to 0.4% for people under 65 [31]. While three equine vaccines are licensed and available for immediate use in the UK [32], no human WNV vaccine currently exists [33], highlighting the need for UK preparedness against the potential health impacts of WNV emergence and transmission on the population.

Risk analysis is a vital element of preparedness planning within both human and veterinary health sectors. In Europe, numerous risk assessments have been conducted to identify the drivers of WNV transmission and to predict regions where cases are most likely to occur [21,34,35,36,37,38,39,40,41]. Similarly, a select number of USUV models have also been developed [42]. However, these analyses were undertaken in countries with documented outbreaks, allowing training of a model and/or validation of the results with observed cases. In this way, the UK presents a challenge, as risk analysis must be conducted in advance of any WNV outbreak.

To date, only one WNV transmission risk analysis for the UK has been undertaken which used a correlative modelling approach to predict current and future WNV risk based on the predicted presence of vectors *Cx. pipiens* and *Cx. modestus*, and presence of corvid and non-corvid hosts [43]. This study showed the greatest WNV risk was in the south-east England, around Greater London, similar to what has been observed with USUV bird cases in recent years. While this study presents a useful foundation for WNV preparedness and surveillance, it can be further complemented by modelling a temperature dependent version of the basic reproduction number, *R*_0_. This metric has been shown in previous studies to be a reliable relative measure of establishment risk of a pathogen [36,44,45,46], as it mechanistically determines whether a complete vector-host transmission cycle can occur; a fundamental requirement for a vector-borne pathogen to sustain itself.

In our study, we extend the approach of Di Pol *et al.* (2022) [36] to England and Wales, building on our previous modelling of current and future distributions of *Cx. pipiens pipiens*, *Cx. pipiens molestus,* and *Cx. torrentium* [47] from a nationwide active mosquito surveillance project [48], thereby incorporating more ecological conditions than temperature alone (for example, precipitation, wind, building density and bioclimatic variables). We also develop the first USUV model-based transmission risk analysis for England and Wales, and compare WNV and USUV risk under present and future conditions. This is particularly important, as the current climate is considered unfavourable for vector competence of WNV, which is greatly reduced under 18°C, but projected changes in temperature and rainfall are likely to alter that [49]. According to the Met Office, the UK has been warming at a rate of approximately 0.25°C per decade since the 1980s, however the most recent decade has been almost double that (0.41°C) [50] which may facilitate autochthonous UK WNV transmission if it continues.

Accordingly, this study provides the first temperature-dependent R₀-based assessment of WNV and USUV transmission suitability in England and Wales, integrating vector biotype distributions and future climate projections. The results are intended to inform surveillance strategies and strengthen disease preparedness in England and Wales, ensuring that public health and veterinary systems are better equipped to respond to future threats from these viruses.

## Methodology

### Suitability metric

The basic reproduction number (*R*_0_) is an epidemiological metric that represents the average number of secondary infections generated by a single primary case in a susceptible population [51]. Here, we used this metric to assess the current and future transmission potential of WNV and USUV in England and Wales. Although USUV was originally detected in the southeast of England, it is becoming increasingly widespread, however transmission and associated disease in wild birds is believed to be only permissive in the summer [10] and therefore we developed risk maps for the most likely period of transmission in the UK which covers July.

In this study, we derive from *R*_0_, presented by Dietz (1993) [52], a suitability metric (*S*), using a similar methodology presented by Di Pol *et al.* (2022) [36] where *R*_0_ is scaled between 0 and 1 to give a measure of relative transmission compared to other regions of the country. Normalisation was undertaken because *R*_0_ is an absolute measure of transmission, but a true estimation is constrained by uncertainties in mosquito and bird density data.

We further extend the *R*_0_ equation into a multi-species equation, where mosquito species *i* is either *Cx. p. pipiens* (*i* = 1), *Cx. p. molestus* (*i* = 2) or *Cx. torrentium* (*i* = 3), and have the species-specific transmission contributions *C_i_*:

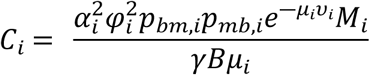

Where *α* represents the temperature-dependent mosquito biting rate, and *α* = 0 for *T* < 3℃ [53], *φ* represents the probability that the mosquito takes a blood meal from a bird, *p_bm_* represents the probability of transmission of the virus from a bird to a mosquito, *p_mb_* represents the probability of transmission from a mosquito to a bird, *μ* represents the temperature-dependent mosquito mortality rate, *υ* represents the temperature-dependent extrinsic incubation period, *γ* represents the recovery rate which is the reciprocal of the infectious period, *M* and *B* represent the mosquito and bird densities respectively. The component *e*^―*μυ*^ represents the probability a mosquito will survive the extrinsic incubation period [54].

We assume that the transmission potential of each mosquito species is independent of each other, and as a result the species-specific contributions add linearly at the level of the total transmission term, giving:

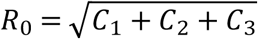

Finally, the normalised value *S*, for location *j*, for disease *z*, is calculated using minimum-maximum scaling, from values derived from all *n* locations across England and Wales (*R*_0, (1)_, *R*_0, (2)_ … *R*_0, (*n*)_) for both diseases, where the minimum value is set to 0, resulting in the ability to interpret *S* = 0 as no risk, and the maximum value is the maximum of all WNV and USUV values combined, to allow for direct comparison between the two diseases:

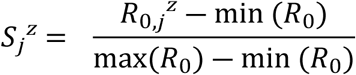

In doing so, we remove the epidemiological threshold where *R*_0_ > 1 represents an outbreak which will take off. An accurate estimation of *R*_0_ is challenging for vector-borne diseases, particularly those transmitted by mosquitoes, as reliable estimates of mosquito density in natural settings are hindered by the challenges of trapping, which captures only a fraction of the population biased by the trapping system, environmental conditions and concurrent host presence. Therefore, the normalised scale can be interpreted as 0 equating to the lowest relative risk (or in our case, no risk), and 1 being the highest relative risk, but should be considered a relative term to the data and parameters used and not necessarily the absolute level of high risk.

Finally, we explore the clinical risk to humans and horses, by multiplying *S* for human (all ages and proportion of over 60’s only) and horse density maps.

### Parameterisation and Host Distributions

The WNV and USUV suitability metrics were parameterised according to Table 1:

**Table 1.** Parameters and equations used in the WNV and USUV suitability metric transmission models and ranges of values identified for the sensitivity analysis.

| Symbol | Description | Species | WNV value | USUV value | Source |
| --- | --- | --- | --- | --- | --- |
| $T_j$ | Temperature (mean of July) for location (j) (degrees Celsius) | NA | 15.59 (11.39 - 23.61) | 15.59 (11.39 - 23.61) | Present: ERA-5 Land 2023 and 2024 average (Muñoz Sabater, 2019) [55]<br>Future: CMIP5 ERA-5 adjusted (Wouters *et al.*, 2021) [56] |
| $\alpha$ | Biting rate per day | All *Culex* | $\frac{q}{43000} T_j (T_j - r)(p - T_j)^{\frac{1}{2}}$<br>$q = 3.71$ (1.86 – 6.17)<br>$r = 1.93$ (–7.93 – 8.74)<br>$p = 45.1$ (36.67 – 59.09) | $\frac{q}{43000} T_j (T_j - r)(p - T_j)^{\frac{1}{2}}$<br>$q = 3.71$ (1.86 – 6.17)<br>$r = 1.93$ (–7.93 – 8.74)<br>$p = 45.1$ (36.67 – 59.09) | Heidecke *et al.*, 2025 [53] |
| $\varphi$ | Biting preference for a bird | *Cx. p. pipiens* | 0.65 (0.45 – 0.85) | 0.65 (0.45 – 0.85) | Griep *et al.*, 2025 (estimated range) [57] |
|  |  | *Cx. p. molestus* | 0.6 (0.4 – 0.8) | 0.6 (0.4 – 0.8) | Griep *et al.*, 2025 (estimated range) [57] |
|  |  | *Cx. torrentium* | 0.5 (0.3 – 0.7) | 0.5 (0.3 – 0.7) | Wehmeyer *et al.*, 2024 (estimated range) [58] |
| $p_{bm}$ | Transmission rate from bird to mosquito | *Passeriformes* to *Culex* | $\frac{1}{(1 + e^{-(bT+c)})}$<br>$b = 0.27$ (0.24 – 0.31)<br>$c = -4.76$ (–5.4 – –4.13) | 0.125 (0.125 – 0.16) | WNV = Heidecke *et al.*, 2025 [53]<br>USUV = Rubel *et al.*, 2008 [59] |
| $p_{mb}$ | Transmission rate from mosquito to bird | *Culex* to *Passeriformes* | 0.88 (0.088 – 1) | 0.88 (0.8 – 1) | WNV = De Wit *et al.*, 2024 [60]<br>USUV = Rubel *et al.*, 2008 [59] |
| $\mu$ | Mortality rate | All *Culex* | $\frac{1}{(-xT_j + y)}$<br>$x = 4.86$ (3.85 – 5.84)<br>$y = 169.8$ (142.1 – 195.6) | $\frac{1}{(-xT_j + y)}$<br>$x = 4.86$ (3.85 – 5.84)<br>$y = 169.8$ (142.1 – 195.6) | Shocket *et al.*, 2020 [46] |
| $v$ | Extrinsic incubation period in days | All *Culex* | $e^{(-dT_j+f)}$<br>$d = 0.08$ (0.07 – 0.1)<br>$f = 5.36$ (5.08 – 5.65) | 14 (7 – 21) | WNV = Heidecke *et al.*, 2025 [53]<br>USUV = Pilgrim *et al.*, 2024 [61] |
| $\gamma$ | Recovery rate (inverse of infectious period in days) | *Passeriformes* | 0.26 (0.097 – 0.359) | 0.182 (0.09 – 0.37) | WNV = De Wit *et al.*, 2024 [60]<br>USUV = Reiczigel *et al.*, 2010 [62] |

A small number of studies have investigated the competence of *Culex pipiens* biotype *pipiens* [61,63,64,65] and *Culex pipiens* biotype *molestus* for USUV [10,61,63,66,67], mostly with small sample sizes and highly variable results, making the definition of USUV–specific transmission parameters more challenging than for WNV. Some laboratory experiments have suggested that USUV is disseminated to the saliva inefficiently in *Culex pipiens* biotype *pipiens* [10,61,63,65], resulting in parameter estimates that do not align well with the increasing numbers of observed bird cases in England [24]. To address this, in the present study we parameterised USUV transmission parameters (*p_bm_* and *p_mb_*) using values based on a model fitted to real-world observed dead bird cases in Vienna [59] instead of competence studies.

Establishing the extrinsic incubation period (EIP) for USUV is particularly difficult, as disseminated infection reaching mosquito saliva in large enough sample sizes of *Culex pipiens pipiens* mosquitoes has proven challenging to achieve under laboratory conditions [10,61,63,65]. It has previously been suggested that the USUV EIP can be adopted from WNV EIP values [42,59,68], despite no evidence supporting this assumption. However, in the absence of robust laboratory-derived parameters, this approach currently represents the most practical alternative. Empirical data indicate that dissemination of USUV can occur across a range of temperatures (see Table 1), typically around 14 days post–infection [61], with reported ranges between 7 and 21 days [66]. This is likely to be temperature-dependent like most arboviruses, however data are too limited to currently model this association. Therefore, the effect of differing EIPs has been presented in the Supplementary Material. Some variation has also been suggested among different *Culex* biotypes [67], although the limited sample sizes in these studies preclude reliable incorporation of such differences into modelling frameworks.

Pilgrim *et al.* (2024) [61] further examined transmission using mosquito competence studies at varying temperatures and found no significant influence on the EIP. However, the lowest experimental temperature tested was 17 °C, whereas mean July temperatures in 2023 and 2024 in our study area reached as low as 12 °C [55]. It therefore remains uncertain whether USUV transmission dynamics differ at lower temperatures than those tested under laboratory conditions.

Given these data limitations, we recommend interpreting the USUV maps primarily in terms of pattern in risk, rather than directly comparing absolute magnitude with WNV. The spatial patterns are expected to be robust to the underlying parameter uncertainty, since they are driven largely by consistent environmental and vector distribution data; however, absolute parameter values remain less certain for USUV than for WNV, and so magnitude-based comparisons between the two viruses should be treated with caution.

Mosquito density per km² was estimated using our previously developed model [47], which applied a General Linear Mixed Model approach to national active surveillance data collected over two consecutive summers. This surveillance identified *Culex pipiens pipiens*, *Culex pipiens molestus*, and *Culex torrentium*, but not *Culex modestus*, a primary WNV vector and important bridge host in Europe whose range is gradually expanding in southeast England [69]. Given its currently reported distribution and low relative abundance compared with the more widespread species included in this risk model, its absence is unlikely to materially affect the resulting risk maps. Nevertheless, the species may still have local epidemiological significance, and its presence in specific areas could influence transmission risk at finer spatial scales.

Bird densities were derived from the 2007–2009 Breeding Bird Survey (BBS) conducted by the British Trust for Ornithology (BTO) [70], which modelled the abundance of 49 species across the UK at a resolution of number of birds per km². This was the most recent modelled bird distribution available for Great Britain. As *Passeriformes* serve as the principal amplifying hosts for both WNV and USUV [15], a density map restricted to Eurasian Blackbird (*Turdus merula*), European Robin (*Erithacus rubecula*), House sparrow (*Passer domesticus*), Song thrush (*Turdus philomelos*)*, Eurasian* Magpie (*Pica pica*), Eurasian Jay (*Garrulus glandarius*), Carrion crow (*Corvus corone*), Common whitethroat (*Curruca communis*), and Garden warbler (*Sylvia borin*), was subsequently developed. These were most likely modelled species to be involved in USUV and WNV long distance movement, during migration, and local maintenance. An analysis was undertaken to explore differences in parameterisation of probability of transmission between bird species, identified in a literature review by de Wit *et al.* (2024) [60], and no statistical significance was observed. Therefore, the nine species of birds included in the model were considered of equal probability to transmit WNV and USUV. However, this is a simplified assumption, as the bird populations are not age-stratified in this model, and nestlings have less feather coverage and defences against mosquitoes and therefore may be more susceptible [71].

At present, the United Kingdom does not have a tracing system in place to know where horses are located. Previously Lo Iacono *et al.* (2012) [72] inferred the spatial distribution of horses by combining land-use data with horse owner records held by the GB National Equine Database (NED). Although there were inaccuracies or gaps in the NED data prior to it being discontinued in 2012 [73], Boden *et al.* (2012) [74] determined the owner location in the NED was similar to horse locations apart from in urban areas such as London. The horse location model by Lo Iacono *et al.* (2012) [72] was cross referenced with locations provided by over 1000 owners and proved to accurately estimate the horse distribution. The alternative, Gridded Livestock of the World (GLW) [75], where the horse distribution has not been updated since 2015, is built upon predictors optimised for intensively farmed livestock. For this reason, the dataset developed by Lo Iacono *et al.* (2012) [72] was used in this study for identifying the horse distribution, and aggregated to a 5km^2^ grid.

Human distributions for 2020 (the most recent available) were downloaded for the all ages and over 60-year-olds in the United Kingdom [76] and rasterised to a 1km^2^ density. For both the horse and human density maps, to reduce local discontinuities and produce a continuous surface, values were smoothed using a multilevel B-spline from the *MBA* package in R-cran [77].

### Sensitivity Analysis

A sensitivity analysis was undertaken using a Latin hypercube sampling partial rank correlation coefficient methodology (LHS-PRCC) [78], where a PRCC value of 0 indicates the parameter is not correlated with the model output, and values of 1 and −1 indicate perfect positive or negative correlation, respectively. The results of sensitivity analyses reflect how parameters behave in the model structure, and not necessarily biological causation. Here, we perform the calculation using 10,000 different parameter combinations from 14 parameters for WNV, and 12 parameters for USUV, plus five additional parameter ranges resembling the three mosquito species abundances, bird abundance and temperature. This was undertaken using the R-packages *lhs* and *epiR.* Parameters were selected using a Latin hypercube sampling method from uniform distributions identified by the ranges in Table 1. The transmission rate of WNV was indeterminate when the infectious period was 0 days, as it resulted in a division by 0, and therefore the 10^th^ to the 90^th^ percentiles of the data were taken.

## Results

### Transmission suitability risk maps

Transmission suitability maps were developed for WNV and USUV across England and Wales for present day (Figure 1) and future (Figures 2 and 3) scenarios, using predicted present and future *Culex* abundance maps from our previous work [47] and parameter values from the sources in Table 1. The maps in Figure 1 are scaled using the maximum value of the maps as 1, whereas the maps in Figures 2 and 3 scaled using the overall maximum value of the future maps as 1. Different scales were used to aid interpretability of the present-day maps.

**Figure 1.**
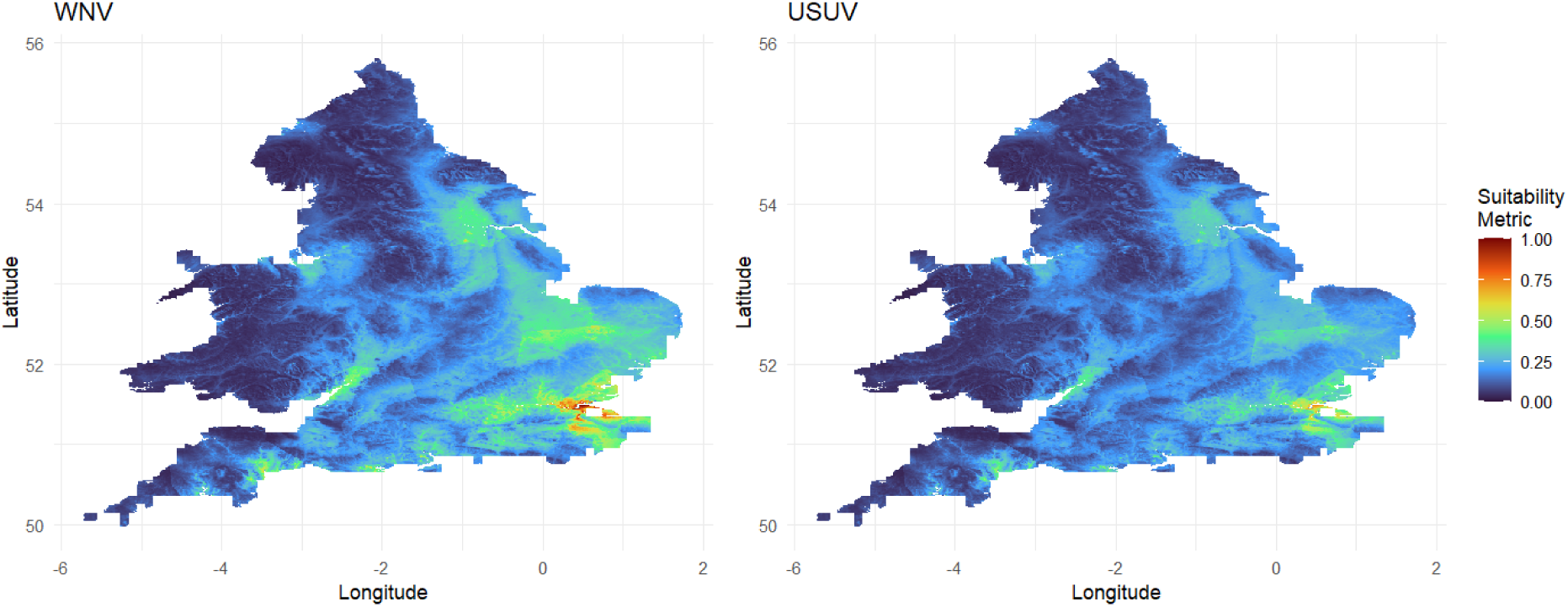
Suitability metric maps for WNV and USUV where 0 resembles no risk and 1 resembles the highest relative risk considering the values of both maps. Both maps demonstrated a similar spatial pattern where there was higher risk in the southeast and east of England, and lower risk in the north and west of England and Wales.

**Figure 2.**
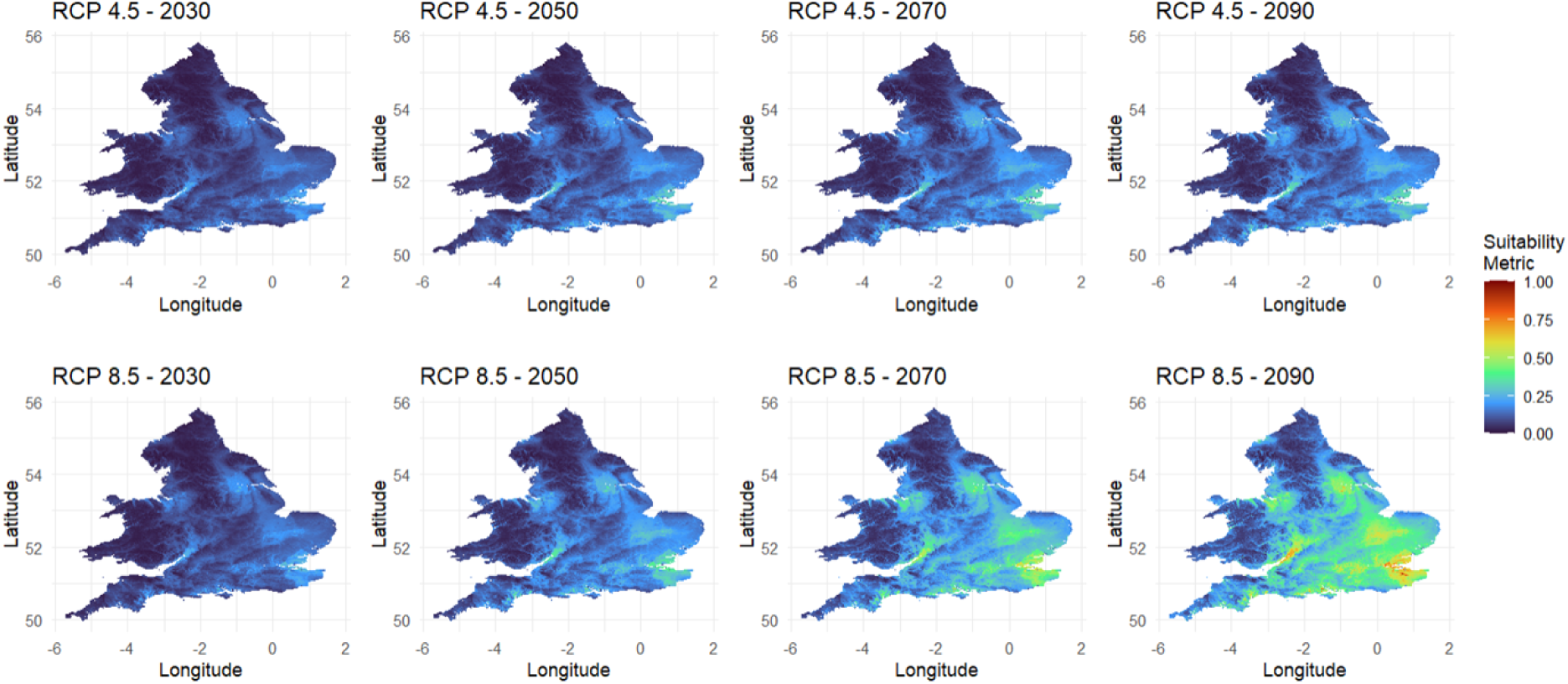
Suitability metric maps for WNV where 0 resembles no risk and 1 resembles the highest relative risk considering the values of all 8 maps. The top row displays changes in risk under future climate pathway RCP 4.5 (moderate emissions) between 2030 and 2090, whereas the bottom pathway displays changes in risk under future climate pathway RCP 8.5 (high emissions).

**Figure 3.**
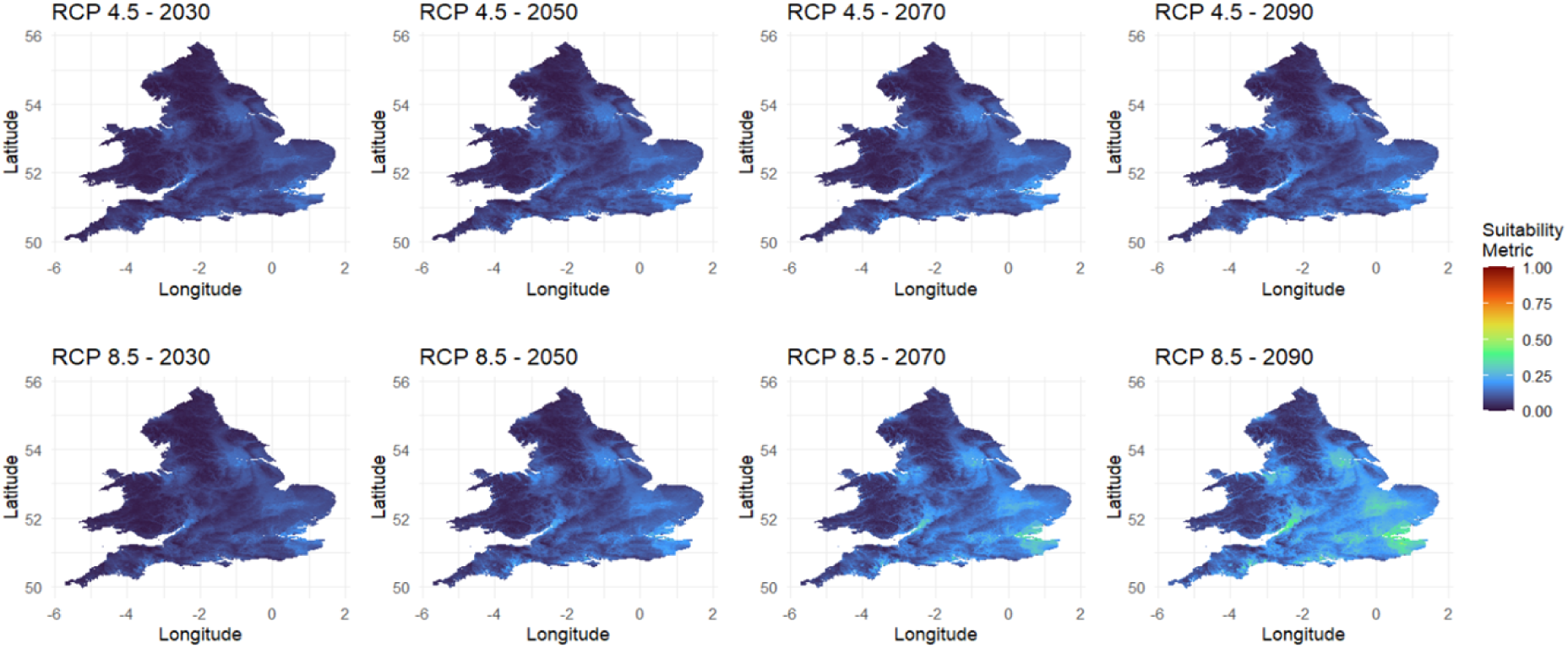
Suitability metric maps for USUV where 0 resembles no risk and 1 resembles the highest relative risk considering the values of all 8 maps. The top row displays changes in risk under future climate pathway RCP 4.5 (moderate emissions) between 2030 and 2090, whereas the bottom pathway displays changes in risk under future climate pathway RCP 8.5 (high emissions).

Using mean temperature data from July 2023 and July 2024 to calculate the temperature dependent parameters gave rise to the suitability metric results in Figure 1. WNV had a higher relative risk than USUV, however as previously mentioned, due to the uncertainty of USUV parameters, the magnitude of risk cannot be reliably compared. Spatially, both viruses displayed a similar distribution showing the highest risk in the southeast along the Thames estuary, spreading into Kent, Essex and Surrey, around the Cambridgeshire and neighbouring counties area, and in the region of Yorkshire, Nottinghamshire and Lincolnshire. This largely followed the spatial distribution of *Culex* mosquito abundance presented in our previous research [47]. Given the uncertainty around the EIP for USUV, further maps were developed for an EIP of 7, 21 and 28 days (Supplementary Figure 1) which displayed very little impact on the level of transmission risk.

The projected future risk of WNV (Figure 2) and USUV (Figure 3) increased over time, with a markedly greater rise under the RCP 8.5 scenario compared to RCP 4.5. This suggests that higher emission trajectories may present climatic conditions more conducive to the transmission of WNV and USUV, likely driven by elevated temperatures and increased mosquito abundance.

Multiplying present-day risk by normalised horse and human densities, or by the proportion of individuals aged over 60 years, produced the clinical exposure risk maps shown in Figure 4. The risk of horse and human cases (the maps on the far left, and middle left) reflects the total risk on the entire population, indicating where the largest numbers of horses or people could be exposed or affected, irrespective of age. In these maps, the areas most likely to experience WNV cases in horses are located in the southeast of England, particularly Newmarket, driven by the high concentration of horses in this region. For humans, London was identified as the area of greatest risk, with Liverpool and Manchester, and to a lesser extent Birmingham, also exhibiting elevated risk.

**Figure 4.**
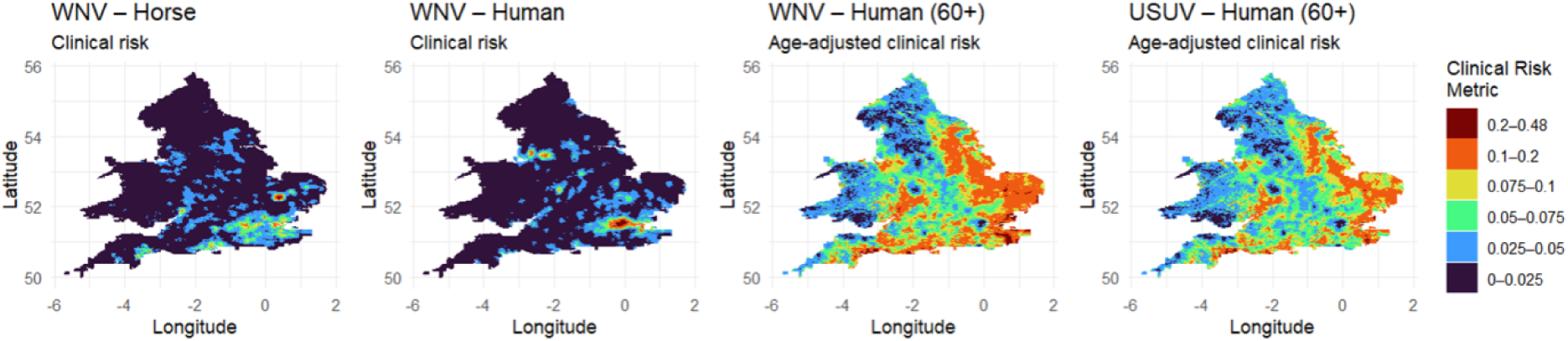
Arbitrary clinical risk of exposure of WNV in the horse (left), human (middle left) and over 60-year-old populations (middle right), and USUV in the over 60-year-old population (right), calculated by the Suitability Metric (on a scale of 0-1) (Figure 1) multiplied by the normalised horse population aggregated on a 5km^2^ grid (on a scale of 0-1), the normalised human 1km^2^ density (on a scale of 0-1), and proportion of over 60-year-olds per 1km^2^ (on a scale of 0-1). Results were categorised to enable visualisation of spatial contrast at lower risk levels.

The maps based on the over 60-year-old proportions represent age-adjusted vulnerability, highlighting areas where higher proportions of older individuals exposed may amplify the likelihood of observing severe clinical manifestations of WNV and USUV in those areas. These maps demonstrate elevated risk in coastal regions, particularly in the east and south of England, with Kent and parts of Norfolk and Suffolk showing the highest risk of observing more severe clinical outcomes.

### Qualitative validation with USUV case data

Positive mosquito and bird USUV cases identified during July and August 2023 and 2024, courtesy of the Vector-Borne RADAR project, were plotted over the present-day USUV suitability map (Figure 5) [24,26]. Cases were visually concentrated in areas of higher predicted suitability, lending support to the spatial risk patterns generated by the model. To complement this visual comparison, we tested statistically whether case locations fell in areas of higher predicted suitability than would be expected under random spatial placement. However, as surveillance effort was targeted rather than spread evenly across the country, and negative surveillance sites were not available for comparison, this test cannot definitively distinguish whether elevated suitability at case sites reflects genuine spatial variation in transmission risk, or simply reflects surveillance being directed toward habitats that also score highly in the suitability model. It should therefore be considered an indicative test of spatial concordance between observed cases and predicted risk, rather than a formal validation of model accuracy. Future work incorporating negative surveillance site data and explicitly accounting for surveillance effort would allow for a more robust assessment of model performance.

**Figure 5.**
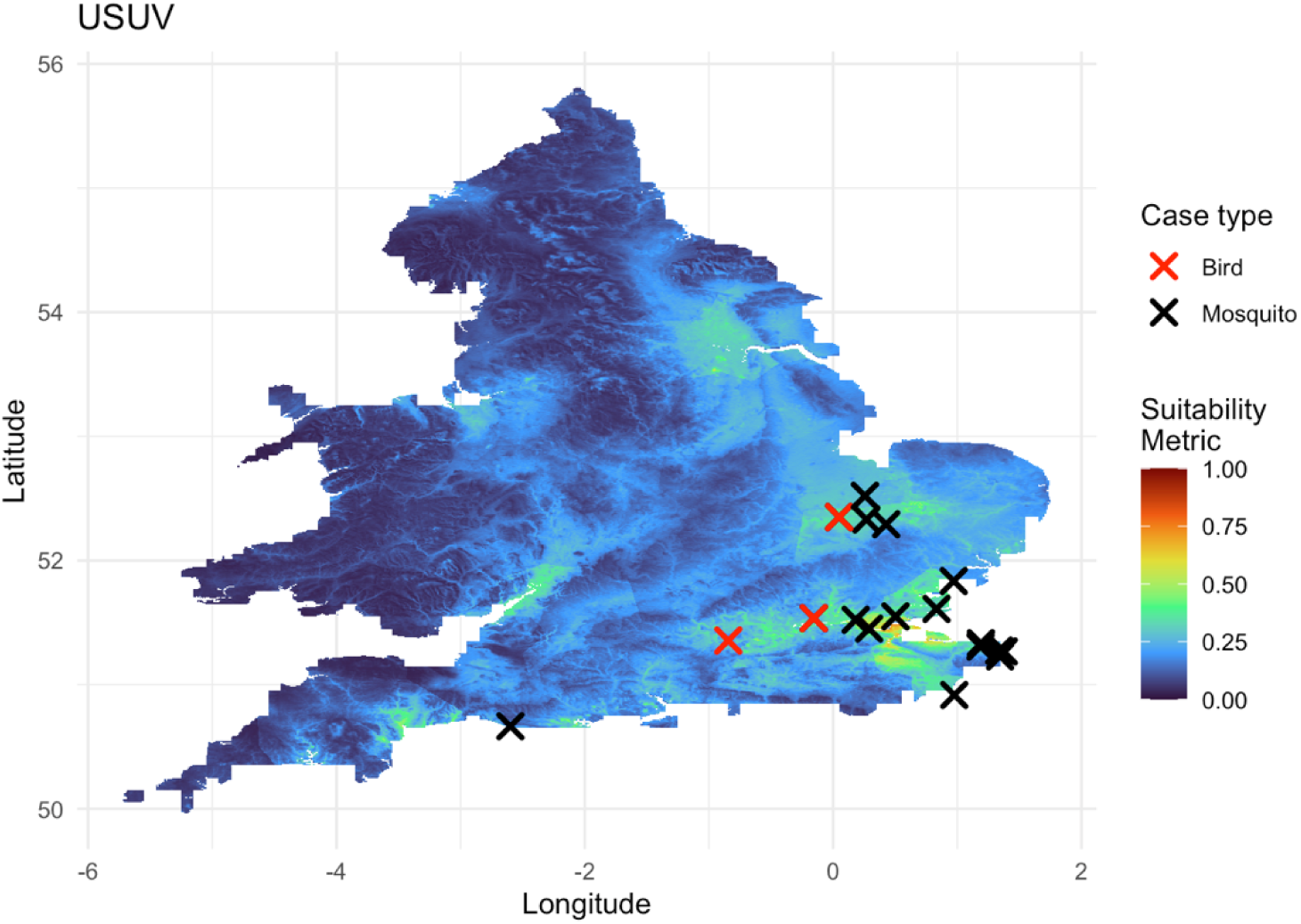
Present day USUV risk map displaying the suitability metric from 0-1, with positive bird and mosquito cases from July and August 2023 and 2024, detected through the VB-RADAR project, overlaid

To assess whether observed USUV cases occurred in areas of higher predicted suitability than expected by chance, we extracted the modelled suitability value at each mosquito case location and calculated the mean suitability across all case sites. We then generated a null distribution by randomly sampling the same number of locations from the suitability raster and calculating their mean suitability, repeating this process 100,000 times. An empirical p-value was calculated as the proportion of random-sample means that equalled or exceeded the observed mean suitability at mosquito case locations (null mean = 0.156, p < 0.001). Bird case locations were not included in the analysis as their flight range was considerably larger than mosquitoes and therefore extracting the suitability metric where they became infected would be less reliable (Figure 6).

**Figure 6.**
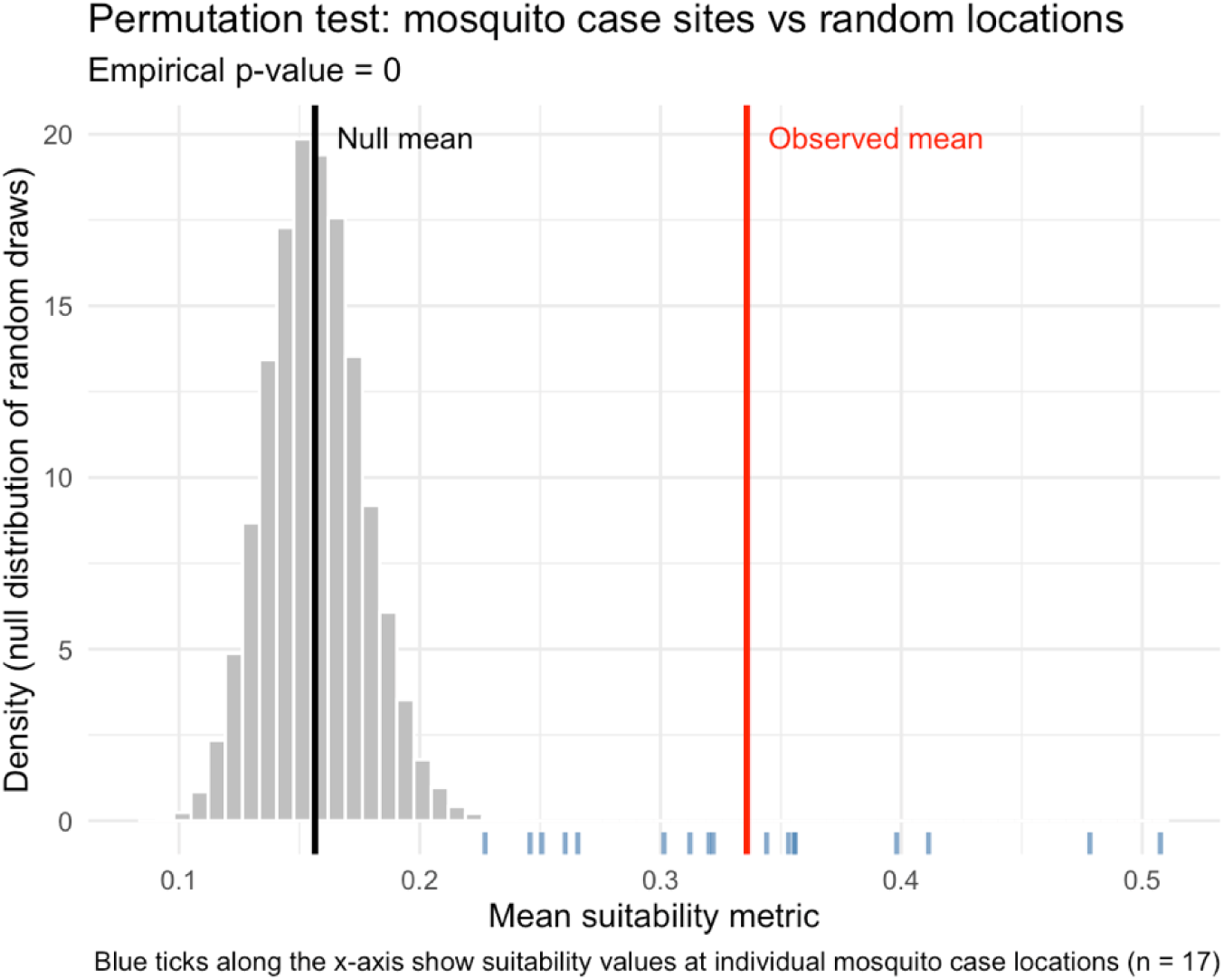
Histogram showing the results of the permutation test. Seventeen random sites were drawn 100,000 times and the mean suitability metric recorded. The null mean (0.156) is the mean of the 100,000 draws. The observed mean (0.336) is the mean of the 17 case locations. Blue ticks resemble the suitability metric in the case locations

**Figure 7.**
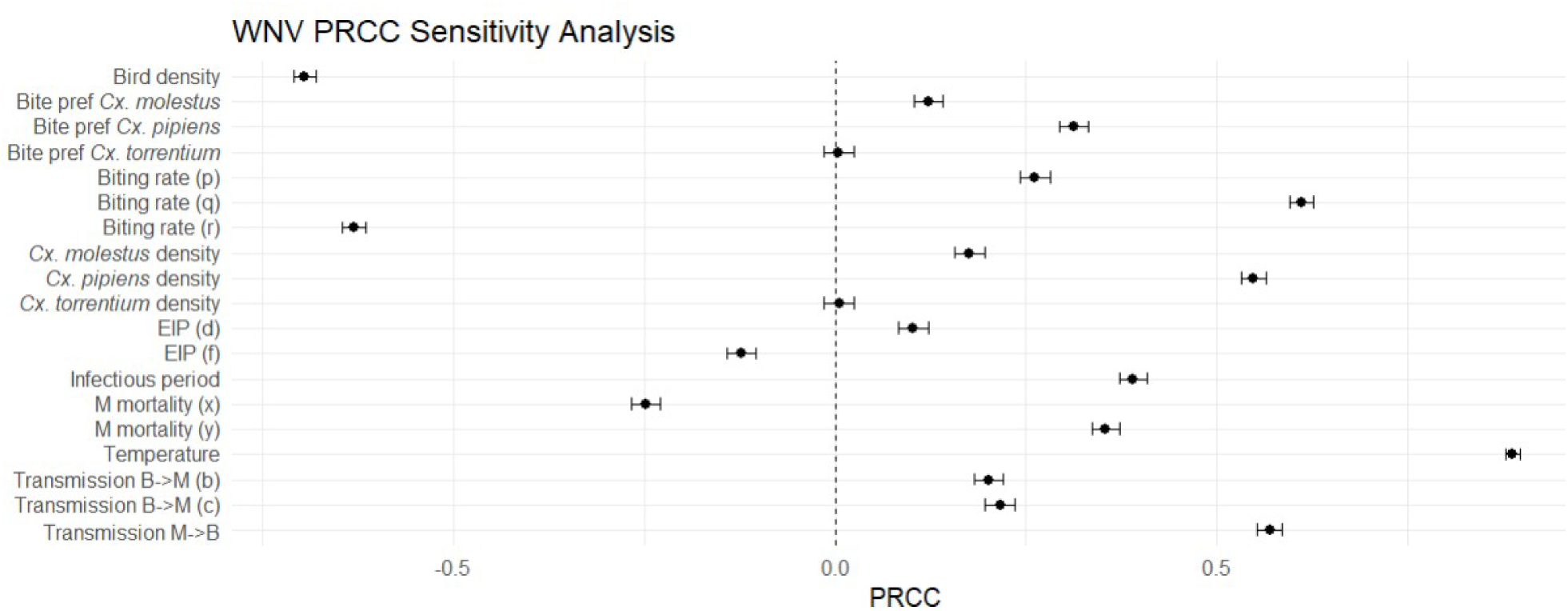
Sensitivity analysis results exploring the parameters in the equation for the WNV suitability metric. A positive or negative PRCC indicates the direction of effect on the suitability metric, the magnitude of sensitivity indicates how important each parameter is in the equation and the confidence interval, represented by the error bars, indicate the uncertainty of the sensitivity analysis.

This permutation-based approach was chosen over a parametric comparison of means as it does not assume normality of the underlying suitability distribution, and directly tests whether the observed spatial pattern of mosquito cases differs from that expected under random placement across the study area. Because case locations arise from targeted rather than randomised surveillance, a significant result indicates that surveyed positive cases were concentrated in higher-risk areas relative to the study area as a whole, but does not on its own establish that this pattern reflects underlying transmission risk rather than the spatial distribution of surveillance effort. With this caveat in mind, the results provide quantitative support for the qualitative pattern described above, indicating that observed USUV cases were not randomly distributed with respect to predicted suitability.

### Sensitivity Analysis

PRCC results for WNV (Figure 5) and USUV (Supplementary Figure 2) were very similar, with near-identical outcomes across most parameters, apart from four. For WNV, the extrinsic incubation period (EIP) was modelled as a temperature-dependent equation in which the two coefficients (*d* and *f*) acted in opposite directions, though each exerted only minor influence on the overall metric. In contrast, the EIP for USUV was not temperature dependent and contributed a small negative effect on the suitability metric. The infectious period exerted a moderate positive influence on the WNV suitability metric, while for USUV the effect was stronger, also in a positive direction. Transmission from bird to mosquito in the WNV model was also modelled by a temperature-dependent equation, with both coefficients (*b* and *c*) yielding small positive contributions in the sensitivity analysis. For USUV, this parameter was constant, producing a positive effect only marginally above zero. Finally, transmission from mosquito to bird had a moderate positive impact on WNV suitability, whereas for USUV the effect was minimal, again only slightly above zero.

In both the WNV and USUV sensitivity analyses, temperature exerted a strong positive influence on the suitability metric, followed closely by the biting rate coefficient (*q*) and *Culex pipiens* density. Bird density and the second biting rate coefficient (*r*) produced the strongest negative effects in both analyses, however bird density in the denominator can produce counterintuitive negative PRCC relationships and therefore must only be interpreted in relation to mosquito abundance. By contrast, *Culex torrentium* density and biting preference had negligible impact on WNV or USUV suitability. Confidence intervals were consistent across the two analyses.

## Discussion

The suitability risk maps presented in this study illustrate spatial variation in WNV and USUV risk across England, with elevated risk in the south and east, and comparatively lower risk in the northwest of England and Wales. Risks to animal and public health associated with both viruses are projected to increase in the future under a changing climate, raising particular concern if WNV emerges, establishes transmission cycles, or overwinters in the UK. Our findings underscore the importance of preparedness for potential WNV emergence, including the development of public education initiatives, ongoing active surveillance in mosquitoes, birds and blood donors, strengthened public health infrastructure, and public health readiness. Given that the emergence of USUV has historically preceded the establishment of WNV in continental Europe, heightened continued vigilance in the UK is warranted to anticipate and respond to potential early WNV incursions, prior to human transmission.

The sensitivity analyses identified very similar PRCC outcomes for WNV and USUV in our study, which indicated that ecological and entomological drivers, such as temperature, biting rate, and *Culex pipiens* density, are central to arbovirus transmission suitability in the England and Wales in general. This highlights that there are common pathways for surveillance and intervention. In particular, the sensitivity analyses revealed strong positive influences of temperature on the suitability of both viruses. This underscores the vulnerability of the UK to climate change and extreme weather event facilitated pathogen emergence events, and the potential for temperature to accelerate key transmission parameters, reducing extrinsic incubation periods and increasing biting rates. But the relationship between temperature and mosquito ecology is likely multifaceted; while higher temperatures may increase biting activity and development of larvae, they can simultaneously reduce the availability of suitable ephemeral aquatic breeding sites [79]. Taken together, these insights suggest that climate-driven suitability is complex, reinforcing the need for ongoing monitoring of mosquito-climate interactions as a critical component of preparedness for mosquito-borne diseases.

It is also important to consider that temperature fluctuations are highly seasonal, especially in temperate zones, and establishment of endemic circulation is dependent on the ability of a virus to overwinter, which USUV has already demonstrated in the UK [25]. This study focused specifically on the month of July, when temperatures are at their highest in combination with *Culex* abundance reaching its peak [80], and therefore when WNV transmission is most likely to occur, making it well suited to identifying periods and regions of peak transmission risk. Overwintering and establishment, however, are governed by a different set of processes, and addressing them would require extending the temporal scope to explore which regions exhibit higher-risk suitability across multiple months of the year, similar to the methodology presented by Di Pol et al. (2022) [36]. Such an analysis would also allow seasonal patterns in virus introduction and transmission suitability to be considered simultaneously. Although most summer bird migrants arrive in the UK between March and May [81], WNV amplification in continental Europe typically occurs later in the summer, meaning many spring migrants are unlikely to be viraemic upon arrival and introduction events may not coincide with peak transmission suitability in the UK (July).

Therefore, focusing on the month of peak suitability may not fully capture the period of public health relevance. Immature *Culex* life stages have been shown to peak in late June, approximately a month ahead of the adult population peak in late July [80], suggesting that a substantial vector population is already established before suitability reaches its seasonal optimum. Should infection be introduced into this earlier population, for instance via late-arriving viraemic spring migrants, low-level amplification could plausibly occur through June, ahead of the peak identified here. Rising temperatures, increased outdoor activity and reduced protective coverage of clothing from early summer onward may further elevate human-vector contact independently of suitability itself. Extending the temporal scope of the model, as discussed already, would therefore allow this earlier period to be assessed directly.

Another aspect to consider, when it comes to public health preparedness, is that areas of higher transmission or clinical case risk do not necessarily correlate with areas of incursion. To understand this combined risk and therefore prioritise surveillance activities and public health resources, transmission risk maps and introduction risk maps need to be combined. At present, two studies have explored introduction of WNV into Great Britain which could provide a foundation for this further work [17,82]. Bessel *et al.* (2016) [17] quantified the risk of introduction of West Nile virus into Great Britain by migrating passerine birds and identified that the area of highest probability for introduction was the southeast and central south coast of England, as far north as Maidstone and Guildford, with a relative moderate probability further north as far as Birmingham. When combined with mosquito abundance predictions [47], hotspots for introduction were limited to coastal regions, with also a slight increase in risk in Cambridgeshire. Brown *et al.* (2012) [82] explored introduction of infected mosquitoes into the UK and identified that Heathrow airport could also be a hypothetical area where infected mosquitoes from aircraft may be introduced, and when overlaid on a score map derived from mosquito and bird abundance, the location of it satisfied the criteria required for potential incursion of WNV.

Ultimately, when combining these incursion studies with the risk maps in our study, three areas become apparent where veterinary and public health resources should initially be focussed: the Kent and Sussex coastline, where increased proportions of individuals aged ≥60 years reside; London, where there is a high density of people and higher temperatures; and Cambridgeshire and surrounding areas including Newmarket, where the greatest concentration of horses in England are stabled. Although our modelling focused exclusively on the *Culex pipiens* group and *Culex torrentium*, the recent expansion of the competent bridge vector *Culex modestus* into Kent, Sussex and Cambridgeshire [69] suggests that local transmission risk in these regions may be further influenced by the presence of this additional vector.

This study has presented the first R_0_-derived risk maps using the results of a nationwide mosquito surveillance study in England and Wales, providing novel insights into both clinical and transmission risk for WNV and USUV. By quantifying spatial relative risk, these analyses contribute to a more refined understanding of where arbovirus emergence and transmission is most likely to occur and may inform decision making for resource allocation, targeted surveillance, and public health preparedness, as well as help to support the recently published UKHSA WNV contingency plan to help guide surveillance [83]. Importantly, the findings highlight the need to integrate climate–driven changes and vector ecology into future modelling efforts, as these factors will shape the trajectory of arbovirus suitability in the UK. Continued refinement and temporal expansion of risk maps, coupled with longitudinal surveillance, will be essential to anticipate potential emergence of WNV, strengthen resilience within the National Health Service, and guide proactive interventions aimed at mitigating the impact of mosquito–borne diseases.

## Data Availability

All summarised data is available within the manuscript or supplementary information. The mosquito dataset supporting this study is available in the VectorByte VecDyn repository at https://vectorbyte.crc.nd.edu/vecdyn-detail/27003.

## Acknowledgements

The authors thank all involved funders, participants and institutions for their contribution to this project. In particular, the following contributors to our previous work developing mosquito distribution models which underpin the work presented in this paper: Amirah Haziqah-Rashid, Emma Widlake, Roksana Wilson, Jolanta Tanianis-Hughes, Agata Delnicka, Fatma Bursali, Amy Jealous, Anthony Abbott, Catie Haines, Colin Johnston, Finn Miller, Ken Sherlock, Sara Gandy and Sarah Biddlecombe. In addition, the authors thank the VB-RADAR project (BBSRC grant number: BB/X017990/1) for sharing mosquito and bird case data to complement the project.

